# Transcriptome and microRNAome profiling of human skeletal muscle in pancreatic cancer cachexia

**DOI:** 10.1101/2025.09.02.25334959

**Authors:** Ashok Narasimhan, Xiaoling Zhong, Brittany R. Counts, Andrew Young, Sha Cao, Jun Wan, Sheng Liu, Leonidas G. Koniaris, Teresa A. Zimmers

**Author notes:** Corresponding author: Correspondence: Teresa A. Zimmers, PhD, 3181 S.W. Sam Jackson Park Road Portland, OR 97239-3098, Mail code: CL3TZ.

## Abstract

**Background and Aims:** Over 80% of patients with pancreatic cancer experience cachexia, characterized by severe muscle and fat loss. While all the mechanistic understanding comes from preclinical models, the translatable nature of these findings to humans remains a critical gap due to the limited knowledge of human cachexia biology.

**Methods:** We generated matched gene and microRNA profiles from rectus abdominis muscle of 55 pancreatic ductal adenocarcinoma and 18 control subjects. Differentially expressed genes and microRNAs were identified at 1.5-fold change and p<0.05.

**Results:** Gene expression results revealed a striking sex-specific difference at the expression and pathway levels. In both sexes, co-expression gene network analysis identified more significant modules and hub genes at 1-month of weight loss than the traditionally used six months, suggesting that gene alterations may be more dynamic in the early stages of the disease progression. When comparing hub genes from humans to experimental models of cachexia, genes such as RELA, DDX21, WDR75, PTPN1, and CRIP3 exhibited similar patterns of expression, suggesting their potential role in cachexia. microRNAs also exhibited sex-specific expression. Although several common miRNAs were identified between sexes, their gene targets differed, indicating that microRNAs may regulate gene targets in a sex-specific manner.

**Conclusions:** The dataset can serve as a resource for validating preclinical findings and exploring previously unexplored molecules in cachexia. Future studies will functionally characterize the role of the hub genes and microRNAs in cachexia. This is the first study to identify sex-specific genes and microRNAs from a single cancer type.

## Introduction

Cancer cachexia is characterized by progressive weight loss^1–3^. More than 80% of patients with pancreatic ductal adenocarcinoma (PDAC) are affected by this debilitating condition^4, 5^. Cachexia is a multifactorial syndrome characterized by involuntary loss of muscle and fat, accompanied by adverse drug reactions, decreased survival, and affects the overall quality of life^6^. Apart from the active catabolism in causing persistent weight loss and muscle depletion, it is also associated with the compromised psychosocial well-being of an individual^7^. Substantial progress has been made in developing clinically relevant experimental cachexia models, and new molecular mechanisms, along with potential therapeutic targets, are constantly emerging. ^8–10^. However, the translatable nature of these findings in human cachexia has remained a challenge. ^11^. This can be attributed to several factors, such as (i) the time point at which muscles are collected in humans and mice, (ii) the heterogeneous nature of the disease, and (iii) the lack of comprehensive datasets generated from a single cancer type to compare with experimental models. With a lack of biomarkers and no therapies, cachexia continues to remain an unmet medical need.

Transcriptome profiling of human fat and muscle has provided valuable insights by identifying genes involved in various cachexia pathways ^12^. Studies from our group have shown that similar pathways can govern muscle and adipose tissue wasting, but by activating a different set of genes^12^. While several of these studies contribute to a better understanding of cachexia in humans, the field lacks comprehensive high-throughput datasets needed for refined analyses, such as understanding the sex-specific alterations in cachexia. Sex-specific differences are increasingly being observed in many metabolic diseases, including cachexia and cardiovascular diseases, but have not been comprehensively explored in human cancer cachexia^13,14^. As well, our understanding of the role of regulatory molecules in human cachexia is at best rudimentary. One such class of regulatory molecules explored in this study is the small non-coding RNAs. Although several classes of small non-coding RNAs have been identified, microRNAs (miRNAs) are widely studied. miRNAs are 18-21 nucleotides in length and are considered global regulators of gene expression^15,16^. miRNAs are involved in diverse roles ranging from regulating myogenesis, and muscle hypertrophy, to various signaling pathways in muscle^17^. Dysregulation of miRNAs leading to muscle atrophy has been documented in aging, Duchenne muscular dystrophy, and cachexia ^18–21^. miRNAs have also been identified as prognostic and predictive biomarkers for cachexia ^19^. While using samples from multiple cancer types may enable us to identify miRNAs involved in global muscle wasting, it precludes our understanding of cancer-specific muscle wasting. In addition, the limited sample size prevents us from performing sex-stratified analysis. The current study addresses these knowledge gaps by producing one of the most extensive datasets of skeletal muscle biopsies from a single cancer type, specifically PDAC.

We performed total RNA sequencing for gene expression and small RNA sequencing to generate miRNA profiles from the same individuals. Sex-stratified analysis at both the gene and miRNA levels revealed striking differences in expression profiles. Further, sex-stratified gene network analyses were conducted using the clinical variable of cachexia to identify canonical pathways and network clusters. We utilized a machine learning approach to identify signatures that can predict weight loss. Finally, several unexplored nodal genes identified in humans were expressed across multiple experimental models of cachexia. Additionally, well-known cachexia genes, such as Trim63, IL6R, and STAT3, have been correlated with muscle wasting in humans and mice.

## Methods

### Recruitment of the study participants

The study was conducted under the protocol #1312105608 approved by the Indiana University Institutional Review Board (IRB). The study participants were recruited at the Indiana hospital between 2014-2018. For the accrual of muscle biopsies, written and informed consent was obtained from the participants. The study included 57 patients with PDAC who underwent surgery and 19 non-cancer controls who underwent surgery for non-malignant conditions, including hernia repair. The rectus abdominis muscle was collected during surgery and snap-frozen immediately until further use for RNA and miRNA sequencing.

### Body composition measurements

Body composition measurements were obtained using computed tomography (CT) and analysis was performed using the SliceOMatic software. CT was obtained as part of the standard of care. CT images obtained before the surgery were used for the analysis. We quantified the skeletal muscle and adipose mass surface area (cm2) using the third lumbar vertebrae as a landmark. Skeletal muscle index (SMI) and total adipose index were calculated by normalizing the surface area to stature (cm²/m²). The patient-generated subjective global assessment (PG-SGA) was used to obtain information on weight loss, which was further confirmed by reviewing the medical records. Cancer weight loss grade, an ordinal variable, was calculated by combining the 6-month weight loss history and BMI information as described by Martin et al^3^. One-month weight loss history and grade were also obtained for these study subjects.

### RNA Isolation, library preparation and Sequencing

RNA was isolated from the rectus abdominis muscle using a combination of QIAzol and the miRNeasy kit (217004, Qiagen, Valencia, CA, USA). 10-20 mg of muscle was excised from the biopsy and immediately placed in Trizol reagent, then homogenized using the Bullet Blender Tissue Homogenizer (Next Advance, NY, USA). After homogenization, the lysate was transferred to a new centrifuge tube for RNA extraction using the Qiagen column-based method. Further purification of RNA was performed using RNase-free DNase (Qiagen #79254). The isolated RNA was quantified using Nanodrop, and the integrity of RNA was confirmed using the Agilent 2100 bioanalyzer system.

Libraries were prepared using Clontech SMARTer RNA Pico Kit v2. As 15 samples had an RNA Integrity Number of 4-6, ultra-low Total RNA-Seq was performed, which can capture both protein-coding and non-coding RNAs. However, we restricted our analysis to the protein-coding genes. One hundred million reads were sequenced for each sample. As part of quality control, a Phred quality score was used to measure the sequencing quality, with more than 95% of the reads achieving 99.9% base call accuracy. The reads were aligned to GRCh38 (hg38) and annotated using Gencode Release 38. The raw reads were normalized using the counts per million (CPM) method. CPM data was log-transformed to perform the gene network analysis.

For the miRNA library, reads were first processed by trimming the 3’ adapter and low-quality bases using cutadapt (cutadapt.readthedocs.io/en/stable/guide.html). Reads with no adapter sequence were tallied (no_adapter_reads). Following trimming, the insert sequences and UMI sequences were identified. Reads with less than 16 bp insert sequences (too_short_reads) or less than 10 bp UMI sequences (UMI_defective_reads) were discarded. To annotate the insert sequences, a unique sequence set was created for all read sets/samples in a submitted job. Following this, the sequences were annotated to mature miRNAs (miRbase V21), where up to two mismatches were tolerated using bowtie (bowtiebio.sourceforge.net/index.shtml). Read counts were calculated from the mapping results.

### Differential gene expression and pathway analysis

For both mRNAs (genes) and miRNAs, differential expression analysis was performed using DESeq2 method in Partek Flow (St. Louis, Missouri, USA). For overall gene and miRNA expression between cases and controls, differential expression analysis was performed after adjusting for age and sex. Further, for sex-stratified analysis, males and females were separated and differential expression analysis was adjusted for age alone. Genes/miRNAs with 1.5-fold change and p<0.05 were considered as differentially expressed. Ingenuity pathway analysis (IPA) (http://www.ingenuity.com/index.html) and Genomatix software were used for biological interpretation. For pathway analysis performed using IPA, we selected pathways with a z-score of 1.5 and p <0.05.

### Identification of transcriptional network using Weighted Gene Co-expression Network Analysis

One of the commonly used methods for identifying a network of genes associated with a clinical trait is Weighted Gene Co-expression Network Analysis (WGCNA). From the complete list of profiled genes (18,750), genes with at least one read count in 90% of the samples were used for the analysis (13,903). Sample clustering based on Euclidean distance was calculated to identify outliers. To determine the threshold for network construction, the pickSoftThreshold function was used to identify the optimal power level, which was 7 in our study. Based on the identified power, module detection was performed where the genes were divided into different modules based on a dynamic mixed method of similarity. Next, the module trait relationship was calculated based on correlation between gene modules and clinical information. The traits used in the study were body mass index (BMI), one-month weight loss grade, one-month weight change, 6-month weight loss grade, and 6-month weight change. Modules with p<0.05 were considered for pathway analysis. If the same module was picked across different traits, the interpretation was performed for only one trait to avoid redundancy as common modules had common genes.

### Identification of hub genes and corresponding gene networks

Hub genes are central genes that are highly connected with other genes in a network. In other terms, these may be considered as upstream regulators which has many downstream effectors in a pathway-based analysis. Being the centrally located gene in the network, they are thought to have a critical role in diseases. Genes identified in each module have a gene significance score and a module membership score. For identifying hub genes, genes with a gene significance of >0.2 and a module membership of 0.9 will be considered for further analysis. The genes that satisfied the set criteria were uploaded to Cytoscape ^22^ using the STRING database plugin to generate the gene interactions. We defined a hub gene as a centrally located gene that had at least 10 interacting genes.

### Identification of differentially expressed miRNAs and target identification

miRNAs with a fold change of 1.5 and p< 0.05 were considered as differentially expressed. The same cut-off was applied to identify sex-specific miRNAs. DIANA tools miRPath v3 was used for pathway analysis. Pathways with p<0.05 were considered significant. The miRNA network was constructed using the Cytoscape App.

IPA (microRNA target filter) was used to identify gene targets of differentially expressed miRNAs and to perform pathway analysis. miRNA targets from sex-specific and overall analyses were identified using the corresponding differentially expressed gene expression datasets (1.5-fold change and p < 0.05). Once the targets were mapped, the target genes were subjected to pathway analysis to identify canonical pathways.

### Machine learning Analysis and deconvolution

We investigated the gene variables that are best predictive of the one-month or six-month percentage weight loss in female, male, and all sample groups. Since the candidate gene variables are high-dimensional, we used penalized linear regression for variable screening, as implemented in the R package “SIS”. SIS stands for sure intendance screening ^23^, and the encoded non-convex sparsity penalty “MCP” has been known to be a stable variable selector ^24^. We used the Bayesian information criterion for selecting the regularization parameter. We aimed to identify sex-specific genes and miRNAs that can predict cachexia at six months and as early as one month weight loss.

For deconvolution, log CPM values were used for sex-specific imputation to identify immune cell populations using CIBERSORT and ICTD-an R Shiny app built to identify cell type proportions, such as adipocytes and fibroblasts, along with immune populations.

### Expression of Hub genes and machine learning genes in experimental mouse models of cachexia

To understand the relevance of hub genes and genes identified from machine learning in cachexia, we investigated the expression of these genes in experimental mouse models of cachexia. Specifically, we used datasets generated from pancreatic cancer (GSE123310), colorectal cancer and colorectal cancer with liver metastasis (GSE142455) and lung cancer (GSE114820). Genes with 1.5-fold change and FDR of 0.05 were considered for analysis.

## Results

### Patient demographics and body composition

The demographics for the study participants are presented in Table 1. Age was significantly different between PDAC and control. Body mass index (BMI) was trending towards significance (p = 0.06), with a lower BMI in patients with PDAC compared to control. The six-month percent weight loss (a 9% reduction in PDAC) and weight loss grade were significantly different between the PDAC and control groups. The one-month weight loss grade was significantly different, but the percent weight change was not. Body composition measurements showed a decrease in skeletal muscle index in PDAC relative to controls, although no significant difference was observed between the groups. A similar trend was observed for muscle attenuation and the total adipose index. Sarcopenia status assigned based on Martin et al classification^25^. There was no significant difference between the groups.

**Table 1:**
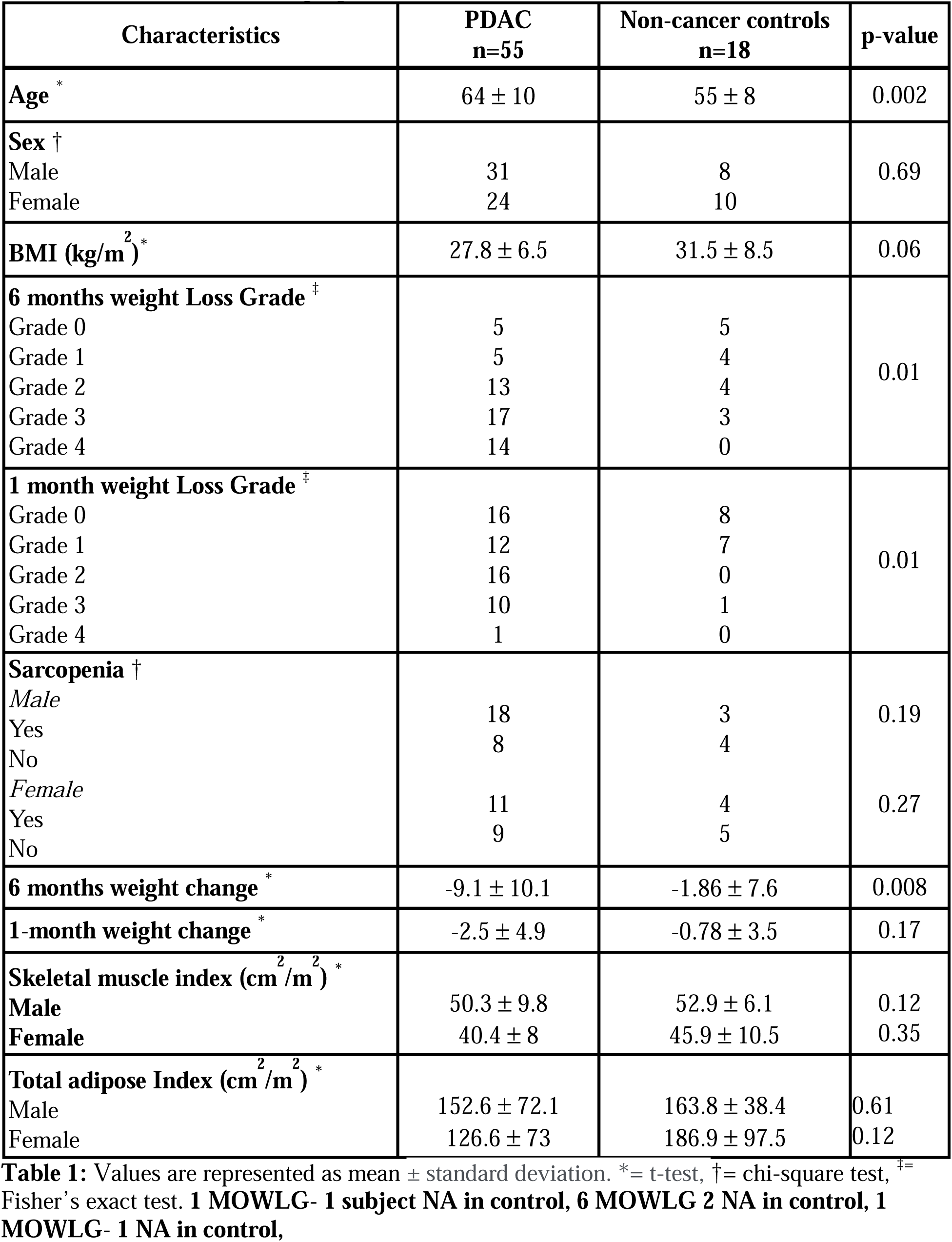
Patient demographics.

### Differential gene expression identified sex specific transcriptome profiles and pathways

We performed differential expression analysis for overall PDAC versus controls (n = 55 vs. n = 18) as well as sex-specific gene expression. There were 31 PDAC and 8 controls in males and 24 PDAC and 10 controls in females. As age was significantly different between the groups and gene expression can be sex-specific, age and sex were adjusted for in the overall analysis, whereas age alone was adjusted for in the sex-stratified analysis. In the overall analysis, 467 genes were differentially expressed at 1.5-fold change and p < 0.05, of which 114 genes were upregulated and 353 were downregulated (Figure 1A). Pathway analysis identified several inflammation-related pathways, including IL-6 signaling, IL-17 signaling, GP6 signaling, and leukocyte extravasation signaling. Signaling pathways, such as the Wnt/β-catenin signaling pathway and the matrix metalloprotease pathway, were also identified (Figure 1B).

**Figure 1:**
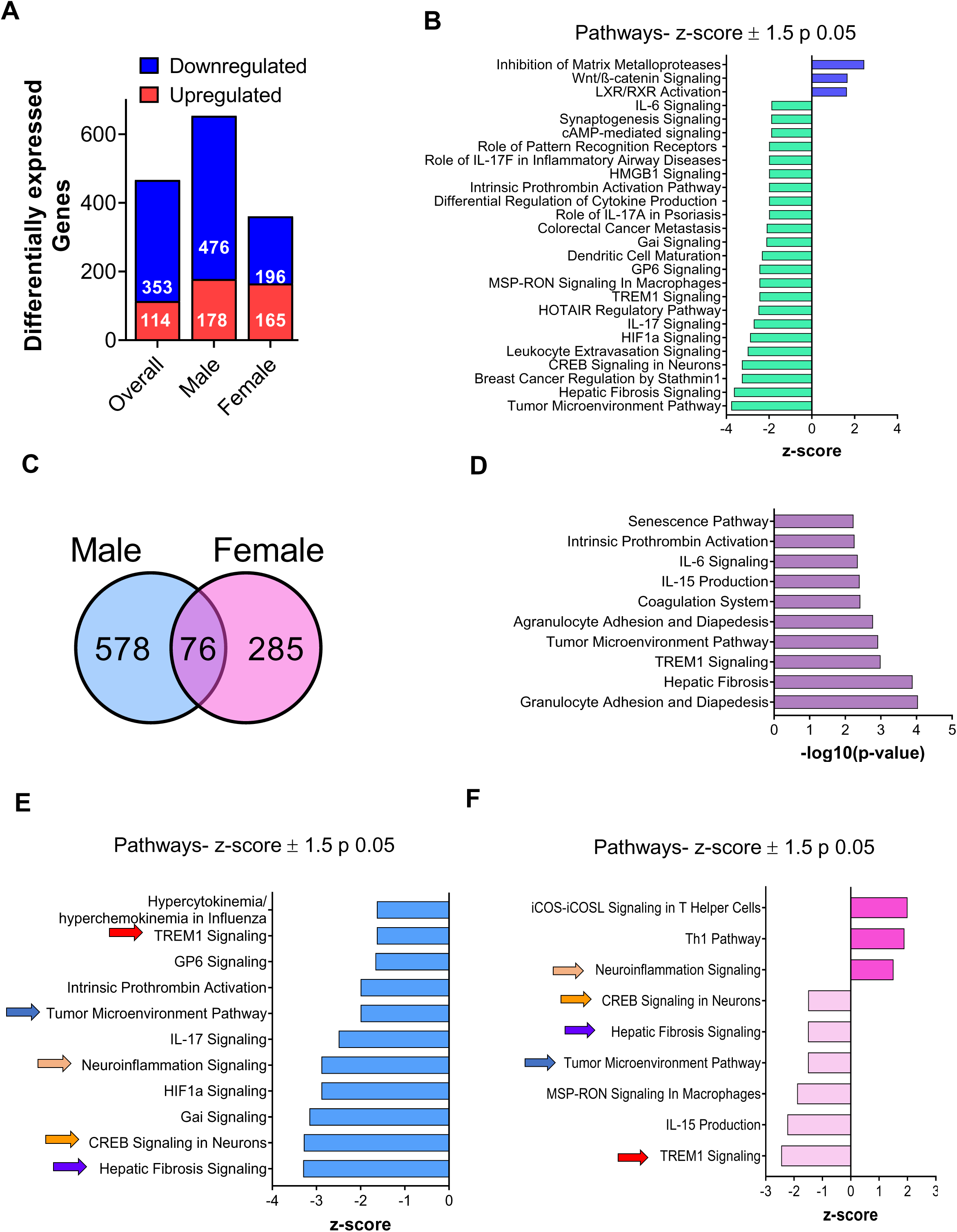
Differentially expressed genes for overall samples PDAC (n=55) vs control (n=18): (A) Age and sex adjusted differentially expressed genes at 1.5-fold change, p<0.05. For the sex-specific analysis (male and female bar graphs), age was adjusted. (B) Pathway analysis for overall samples with z-score ±1.5 and p<0.05 is represented. (C) Differentially expressed genes showed minimal overlap between sexes. (D) Pathways for 76 overlapping genes. Canonical pathways for males (E) using the 578 unique genes and females (F) using 285 unique genes. Common pathways in both sexes were indicated with arrows.

We identified a distinct set of genes for males and females with minimal overlap. In males, 654 genes were differentially expressed, of which 178 were upregulated and 476 were downregulated. In females, 361 genes were differentially expressed – 165 upregulated and 196 downregulated. Seventy-six genes were common between both sexes, of which 67 were expressed in the same direction (Figure 1C). Inflammatory pathways were identified in both males and females, which is a hallmark of cachexia (Figure 1D). Several common and unique pathways were identified in males and females. To determine if the same pathway affects different gene profiles in a sex-specific manner, we focused exclusively on the common pathways. Indeed, TREM-1 signaling, neuroinflammation signaling, and hepatic fibrosis signaling were common between males and females (Figures 1E-F); however, the gene overlap for these pathways was minimal (Supplementary Figures 1A-E). To summarize, our data suggest distinct sex specific muscle transcriptome profiles and pathways are involved in causing muscle wasting in PDAC cachexia.

### Network co-expression analysis identified sex-specific network modules

For WGCNA, PDAC samples alone were considered for network analysis along with the clinical traits. First, we performed the network analysis for all 55 PDAC samples using 6-month weight loss grade and weight change, 1-month weight loss grade and change, BMI, and sex. Modules with p<0.05 were identified as significant modules. If the same module was picked up in two traits, only one module was considered for interpretation to avoid redundancy. Interestingly, we observed a significant number of modules associated with one-month weight change and grade compared to the traditional six-month weight loss and grade (Figure 2A). This suggests that the molecular alteration in muscle is more dynamic at early time points and may plateau over time. We performed pathway analysis for all the standard significant modules identified in BMI, 1– and 6-month weight loss grade, and weight change. Many known pathways, such as cellular response to stress, Hedgehog signaling, IL-6 signaling, PI3K/AKT, mTOR signaling, TGF-β receptor signaling, the ubiquitin proteasome pathway, and apoptosis, were common to all five traits (Figure 2B)^1, 26–28^. Additionally, several novel pathways, including the EGFR1 pathway, PDGFR-beta, TSH signaling, and vesicle-mediated transport, were identified (Figure 2B). When analyzed for unique modules in BMI, one-month weight loss, and grade, several metabolic pathways associated with inflammation and signaling pathways were identified (Figure 2C). In all, several known pathways mediating cachexia, including those involved in inflammation, protein synthesis and degradation, and signaling pathways in muscle wasting, were identified, along with other novel pathways.

**Figure 2:**
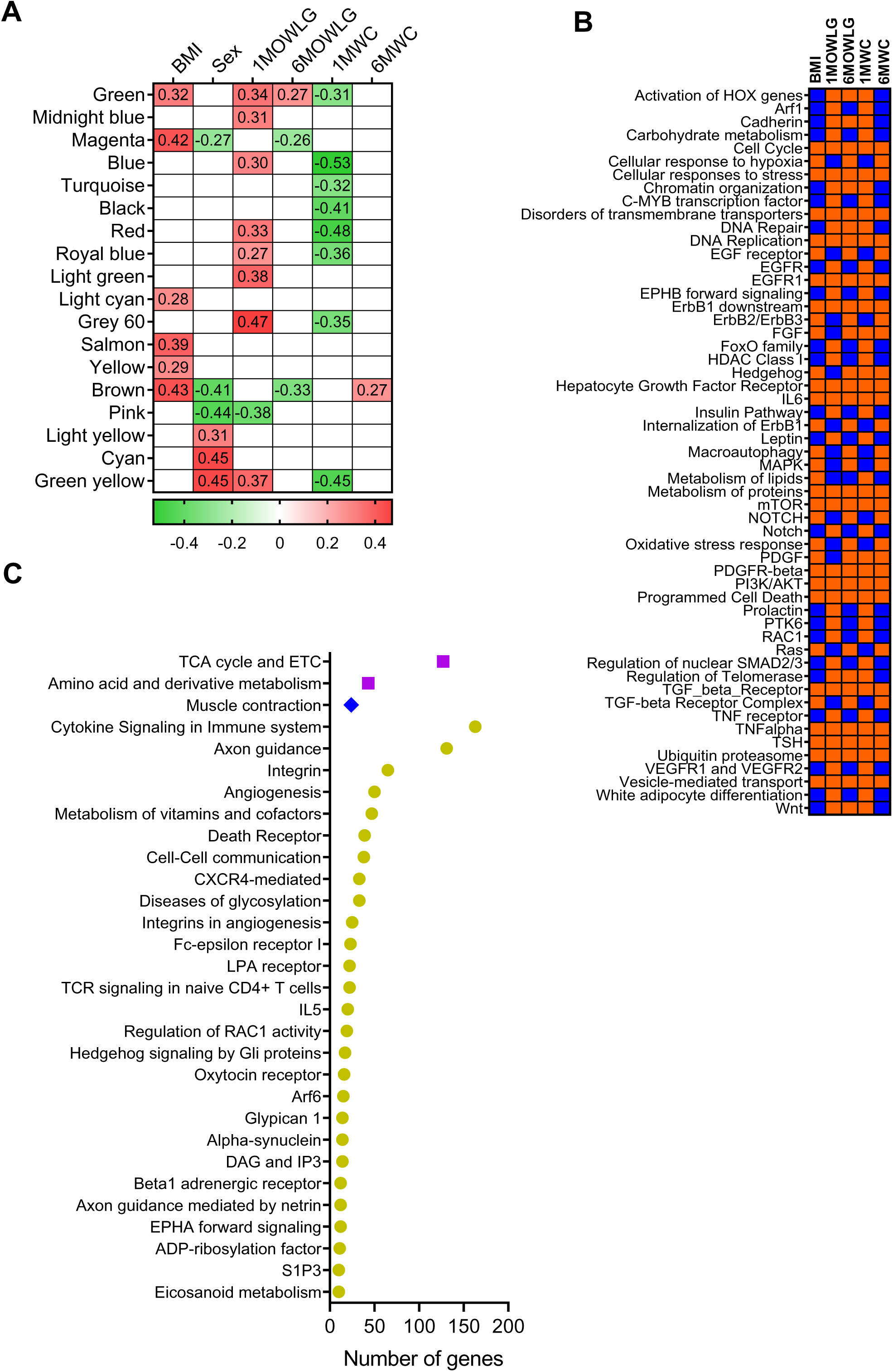
WGCNA for the PDAC samples: (A) Module trait relationship where the correlation values for the significant modules are indicated. (B) Pathway analysis for all common significant modules in all traits. Orange indicates the presence of pathways, and blue indicates the absence of pathways. (C) The dot plot is for the pathways that are unique to one trait: Purple square-BMI (light cyan, salmon, yellow), Blue diamond-1MOWLG (midnight blue, light green), green circle-1MWC (turquoise and black).

In addition, we performed sex-stratified WGCNA using 31 male PDAC and 24 female PDAC samples, as we found many significant modules associated with sex as a variable (Figure 2A) in the overall analysis. Similar to the overall PDAC analysis, many significant modules were identified in one month weight change (Figure 3B). However, none of the modules were significant in the six-month category and the one-month weight grade. The correlation for the significant module is presented in Supplementary Figure 2. Along with identifying the known cachexia pathways, less explored pathways such as Arf6 signaling, sphingosine 1-phosphate pathway, and angiogenesis (Figures 3C-E) were identified. The hub genes identified in males were ARF3, FYN, and PIP4K2A, which are involved in vesicle trafficking, control of cell growth, and cell proliferation, respectively (Figure 3F). The mechanistic roles of these hub genes have not been investigated in cachexia yet.

**Figure 3:**
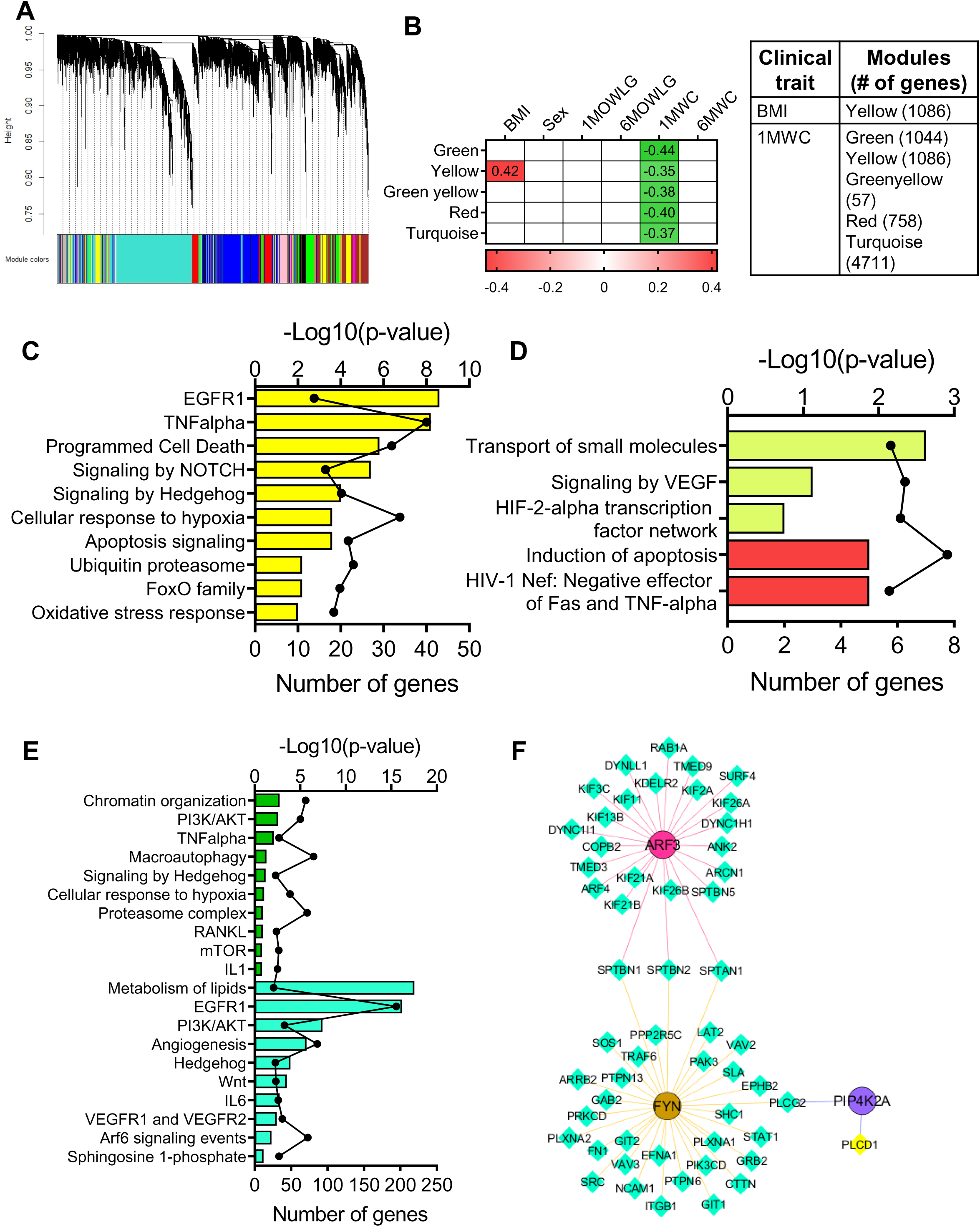
WGCNA for male PDAC samples: (A) Cluster dendrogram representing all the identified modules. (B) Correlation values for the significant modules at p<0.05 along with the number of genes present in each module. (C) Pathway results for the genes present in yellow module for BMI. Pathway results for genes identified in green-yellow and red modules (D), and green and blue modules (E) for 1MWC. (F) Hub genes are circled with different color schemes in the center along with their downstream nodal networks from turquoise module. The color of the downstream indicate the module colors.

Consistent with the overall analysis and male PDAC analysis, eight significant modules were associated with one-month weight change in females. (Figure 4B). Pathways related to metabolism, such as the TCA cycle, the electron transport chain pathway, amino acid metabolism, cell cycle-related pathways, such as the ATM pathway, apoptosis, cell cycle checkpoints, the DNA repair pathway, and inflammatory pathways, such as the innate immune system pathway, TGF-β receptor signaling, were identified from female network modules. Novel pathways in females include VEGFR1 and VEGFR2 signaling and alpha 6 beta 4 pathway (Figures 4C-D). Common pathways between males and females include Hedgehog signaling, PI3K/AKT, EGFR1, ubiquitin proteasome and mTOR signaling. However, the genes involved in activating these common pathways are predominantly unique to each sex, highlighting that distinct transcriptome profiles can activate the same pathway. Several hub genes including STAT5A, RELA, POLDIP3, XRN2, ACTR2, CCT2, CEP97, SEC31A, and COPB2 (Figures 4E-G). The correlation between module membership versus gene significance and significant pathways associated with BMI, six months weight loss, one month weight loss grade is presented in Supplementary Figures 3A-F.

**Figure 4:**
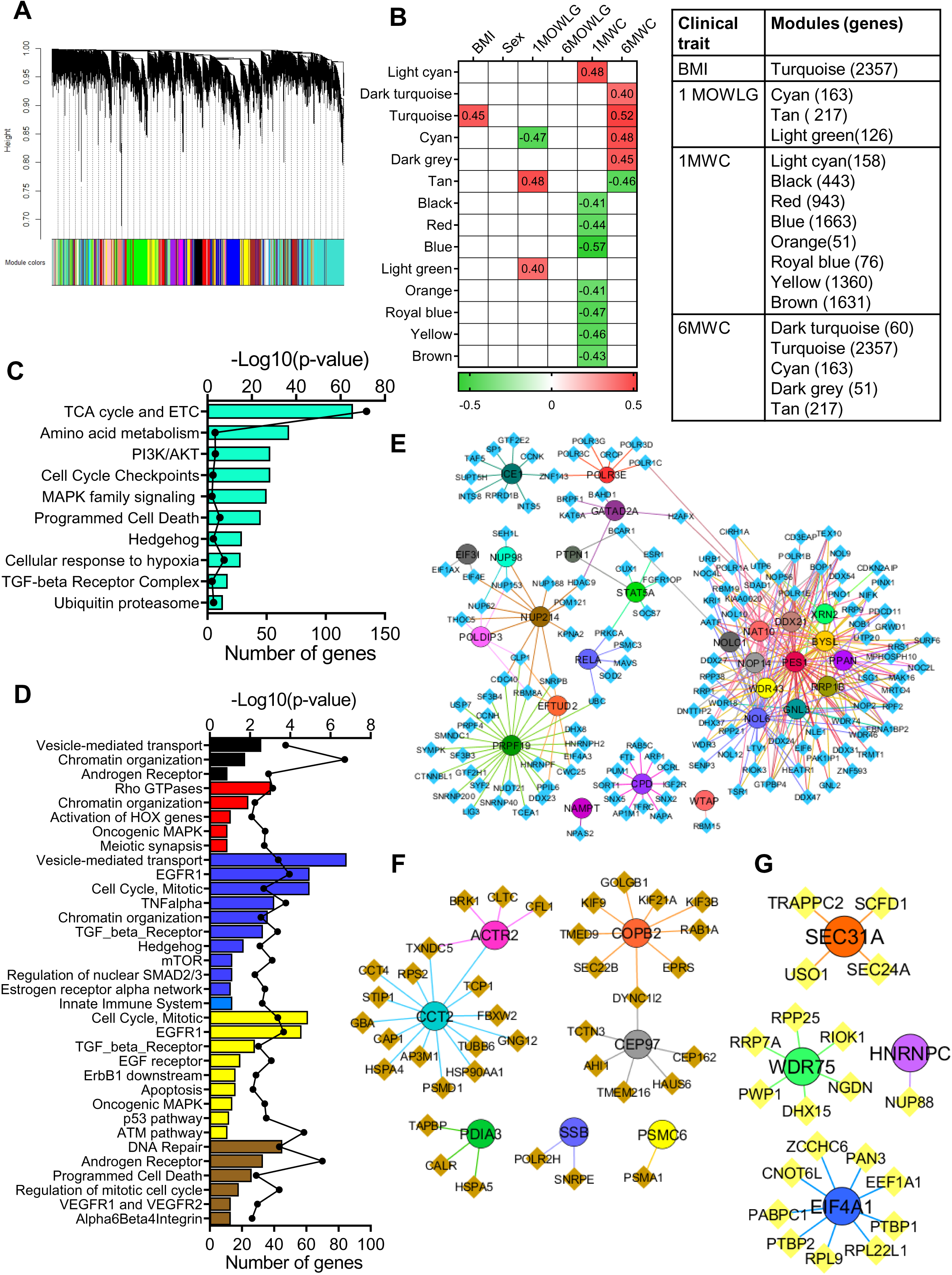
WGCNA for female PDAC: (A) Cluster dendrogram representing all the identified modules. (B) Correlation values for the significant modules at p<0.05, along with the number of genes present in each module. (C) Pathway results for the genes present in the turquoise module for BMI. (D) Representative significant pathways from black, red, royal blue, yellow, and brown modules from 1MWC. (E-G) Hub genes are circled with different color schemes in the center along with their downstream nodal networks. The color of the downstream indicates the module colors.

### Sex specific miRNA profiles and pathways

Dysregulated miRNAs were identified from the same muscle biopsies that were used for gene expression studies after adjusting for age and sex. Since we did not have sufficient RNA from a control sample, 17 control samples were sequenced. 242 differentially expressed miRNAs were identified from the overall PDAC vs control (n=55 vs n=17) (Figure 5A). In the sex stratified analysis, 124 miRNAs and 240 miRNAs were differentially expressed in males and females, respectively. 68 miRNAs were common between both sexes (Figure 5B). The pathways targeted by 68 common miRNAs included IL6 signaling, NRF2-mediated stress, mevalonate pathway, Integrin signaling, and 3-phosphoinositide signaling (Figure 5C). The role of many of these pathways need to be elucidated in muscle wasting. We next wanted to understand the roles of unique miRNAs in males and females. Pathway analysis using the unique miRNAs identified 14 common pathways between males and females (Figure 5D).

**Figure 5:**
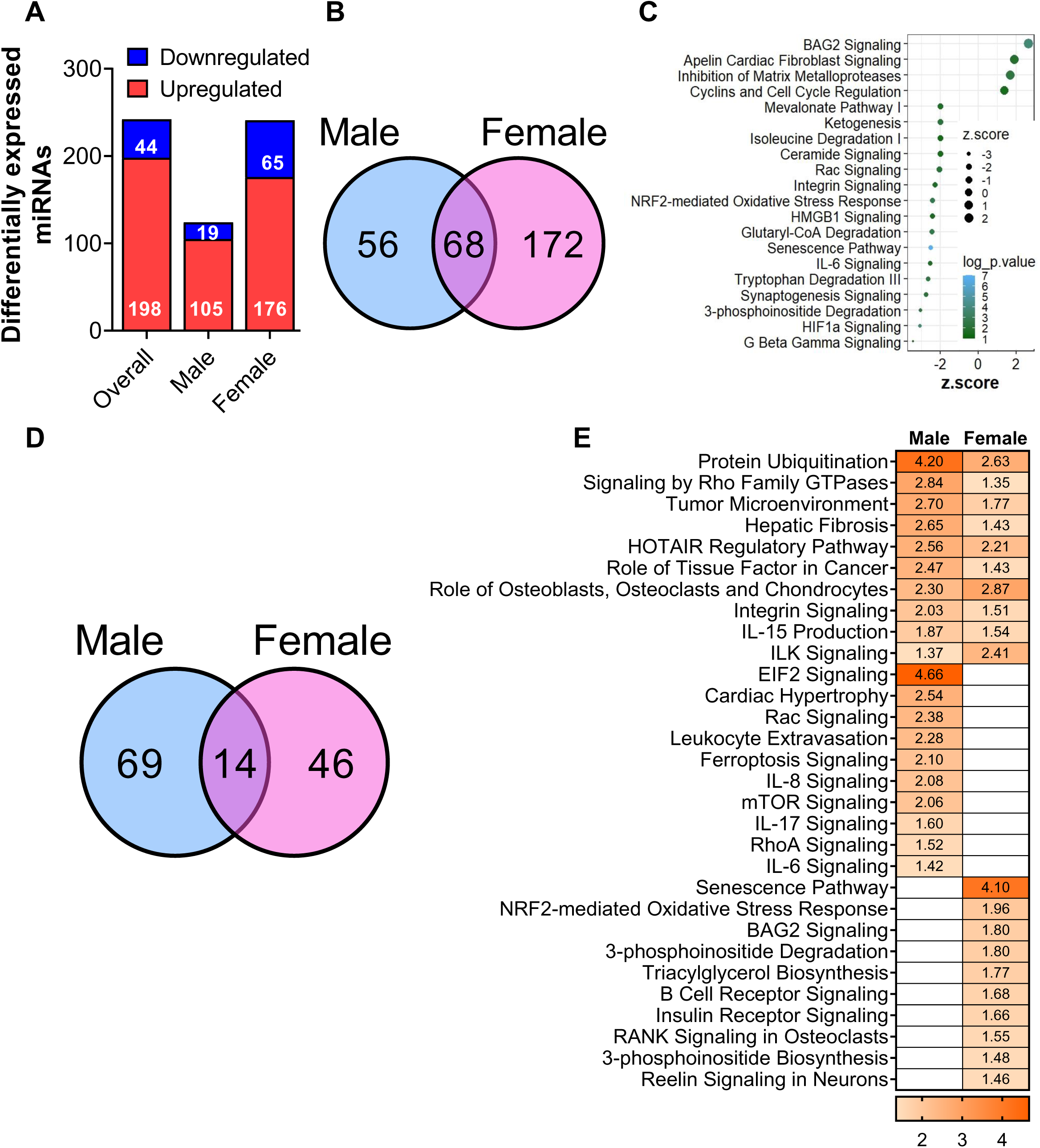
miRNA-differential expression: Differentially expressed genes for overall samples PDAC (n=55) vs control (n=18): (A) Age and sex adjusted differentially expressed genes at 1.5-fold change, p<0.05. For the sex-specific analysis (male and female bar graphs), age was adjusted. (B) Comparison of age-adjusted differentially expressed sex-specific miRNAs between males and females. (C) Representative pathways identified from 68 common miRNAs with the circles and colors corresponding to z-score and log p-value, respectively. (D) Number of pathways identified from the unique miRNAs for males and females. 14 common pathways were identified. (E) 10 representative pathways, each from males, females, and common between them, are presented along with their z-scores.

One of the well-characterized properties of miRNAs is their pleiotropism in regulating several genes ^29^ and their redundant properties where different miRNAs may regulate a single gene ^30^. This phenomenon was observed in our dataset. For example, IL-6 signaling was identified in the common 68 miRNA pathway analysis. Similarly, IL-6 signaling was identified in males when the unique miRNAs were analyzed (Figure 5E). A similar interpretation could be extended to the NRF2-mediated oxidative stress response, which was identified in both common pathways and the female analysis (Figure 5E). Several pathways were found to be unique to males, including mTOR signaling and RhoA signaling. The unique pathways in females included triacylglycerol biosynthesis, 3-phosphoinositide degradation to name a few. These results highlight the complex interaction that could potentially occur between different molecules (miRNAs and genes in this context) in regulating different pathways in a disease specific context.

We next extended our analysis to explore the 14 common pathways. While several unique miRNAs were identified for each pathway, we focused only on the common miRNAs with more than 1.5 fold change for representation (Figures 6A-C). Although the miRNAs that regulate these pathways were common, the mRNA targets were strikingly different. These results emphasize the fact that the same miRNAs can regulate a common pathway in males and females by regulating a different set of genes in a sex-specific manner.

**Figure 6:**
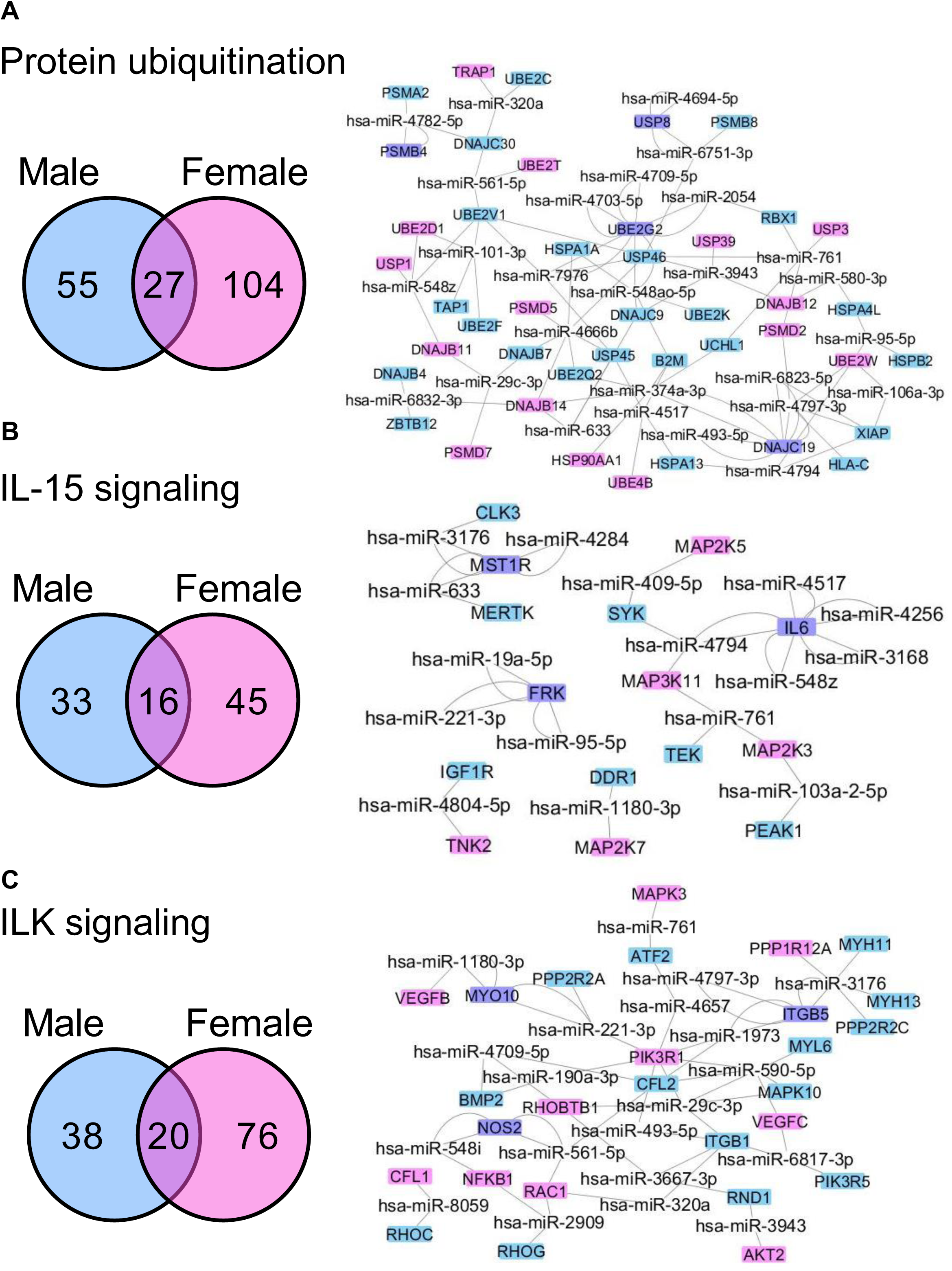
miRNA-mRNA pathways with targets (A-C): Three representative pathways common between males and females are shown with miRNAs and their cognate gene targets. Each pathway had several unique and common miRNAs. Network diagrams represent the gene targets for common miRNAs. Gene targets highlighted in blue indicate male-specific genes, genes highlighted in pink represent female-specific genes, and those highlighted in purple are genes common to both males and females.

### Machine learning analysis identifies sex-specific gene and miRNA signatures

Machine learning analysis identified sex-specific genes to predict cachexia at one month and six-month weight loss stage (Table 2). At one month weight loss, the algorithm identified DOCK9, WDR60, PCSK7, PROSER1, ZNF324, SPTLC2, TCTA, TYW3, VCPKMT in males. ACTR5, CPT1A, FOXJ3, OGA, P4HB AND ZNF354A were identified in females.

**Table 2a:** Machine learning mRNA – sex specific analysis.

| **1 month percentage loss** |  | **6 months percentage loss** |  |
| --- | --- | --- | --- |
| *Male* | *Female* | *Male* | *Female* |
| DOCK9 | ACTR5 | CRIP3 | CCL27 |
| WDR60 | CPT1A | ZSWIM2 | CEP112 |
| PCSK7 | FOXJ3 | KLK10 | GMNN |
| PROSER1 | OGA | LALBA | KCTD9 |
| ZNF324 | P4HB | OR11L1 | METAP2 |
| SPTLC2 | ZNF354A | PMP2 | PARK7 |
| TCTA |  | PTF1A | PSMB5 |
| TYW3 |  | TBC1D3B |  |
| VCPKMT |  |  |  |

**Table 2b:** Machine learning miRNA – sex specific analysis.

| **1 month percentage loss** |  |
| --- | --- |
| *Male* | *Female* |
| hsa-miR-1255b-5p | hsa-miR-128-1-5p |
| hsa-miR-1911-5p | hsa-miR-200b-3p |
| hsa-miR-301a-5p | hsa-miR-34c-3p |
| hsa-miR-3164 | hsa-miR-4511 |
| hsa-miR-511-5p | hsa-miR-4730 |
| hsa-miR-548o-3p | hsa-miR-548ak |
| hsa-miR-599 | hsa-miR-5701 |
| hsa-miR-6124 | hsa-miR-6716-3p |
| hsa-miR-640 | hsa-miR-6814-3p |
| hsa-miR-656-3p | hsa-miR-6830-3p |
| hsa-miR-6737-3p | hsa-miR-765 |
| hsa-miR-676-3p |  |
| hsa-miR-6836-5p |  |
| hsa-miR-6869-3p |  |
| hsa-miR-7110-5p |  |

In six-months weight loss category, the genes identified in males included CRIP3, ZSWIM2, KLK10, LALBA, OR1L1, PMP2, PTF1A AND TBC1D3B. The genes identified in females included CCL27, CEP112, GMNN, KCTD9, METAP2, PARK7 and PSMB5. The functions of all these genes identified in both categories remains unknown in cachexia.

Similar pattern was observed in miRNA analysis. At one month weight loss, the miRNAs identified in males included hsa-miR-125b-5p, hsa-miR-1911-5p, hsa-miR-301a-5p, hsa-miR-3164, hsa-miR-511-5p, hsa-miR-548o-3p, hsa-miR-599 and hsa-miR-6124. The miRNAs identified in females included hsa-miR-128-1-5p, hsa-miR-200b-3p, hsa-miR-34c-3p, hsa-miR-4511, hsa-miR-4730, hsa-miR-548ak, hsa-miR-5701 and hsa-miR-6716-3p.

In six month weight loss category, the miRNAs identified in males included hsa-miR-1273h-3p, hsa-miR-203a-5p, hsa-miR-204-3p, hsa-miR-2276-3p, hsa-miR-3192-3p, hsa-miR-3913-3p, hsa-miR-4417, hsa-miR-4668-3p, hsa-miR-4999-5p, hsa-miR-583, hsa-miR-639, hsa-miR-6760-3p, hsa-miR-6762-3p and hsa-miR-6886-3p. The miRNAs identified in females included hsa-miR-216a-3p, hsa-miR-2355-5p, hsa-miR-4468, hsa-miR-4518, hsa-miR-4792, hsa-miR-5002-5p, hsa-miR-6512-5p, hsa-miR-6741-3p, hsa-miR-6744-3p, hsa-miR-6824-5p, hsa-miR-6887-3p, and hsa-miR-873-3p.

As inflammation is a hallmark of cachexia, we used the deconvolution approach to identify specific immune populations and performed an overall and sex-stratified analysis. In an overall analysis using ICTD, neutrophils and fibroblasts showed significant differences between the PDAC and control groups (Supplementary Figures S4A-B). The proportions of fibroblast and neutrophils were increased in PDAC when compared to controls. In males (Supplementary Figures S4C-D), fibroblasts, plasma cells, T follicular helper cells and TNK cells were significantly different between PDAC and controls. In females (Supplementary Figures S4E-F), M1 macrophages was significantly different between PDAC and controls. These results, although obtained through *in silico* predictions, potentially suggest that inflammation and fibrosis could play a role in muscle wasting. Future studies may utilize single-cell RNA sequencing to specifically define the immune populations that are altered during muscle wasting.

### Potential new molecules for further functional characterization in cachexia identified from hub network and machine learning

The experiment was aimed at identifying potential molecules that can act as common drivers of muscle wasting in cachexia across cancer types. To this end, we used pancreatic cancer, colorectal cancer, and colorectal cancer with liver metastasis datasets ^31–33^. Except in the lung cancer study, where the gastrocnemius muscle was used for sequencing, the remaining three studies used the quadriceps muscle. Hub genes represented in Figures 4 and 5, and machine learning genes identified from one and six-month weight loss changes, were queried in these three datasets. We considered genes that were differentially expressed at a 1.5-fold change and an FDR of 0.05 in all four experimental models. From the hub gene comparison, 13 genes were commonly present across all four cancer types with the same direction of effect (Figure 7A). These genes included NUP98, PSMC6, WDR43, RELA, and SEC31A, among others. From the machine learning studies, in one month weight loss, P4HB was present across four datasets while TYW3, FOXJ3 AND TCTA were common in GSE123310 and GSE142455 (Figure 7B). In six-month weight loss, CRIP3 was the only gene present across all datasets (Figure 7C). Except for RELA ^34^, the role of other genes is not characterized in cachexia serving as potential novel molecules to understand their role in muscle wasting.

**Figure 7:**
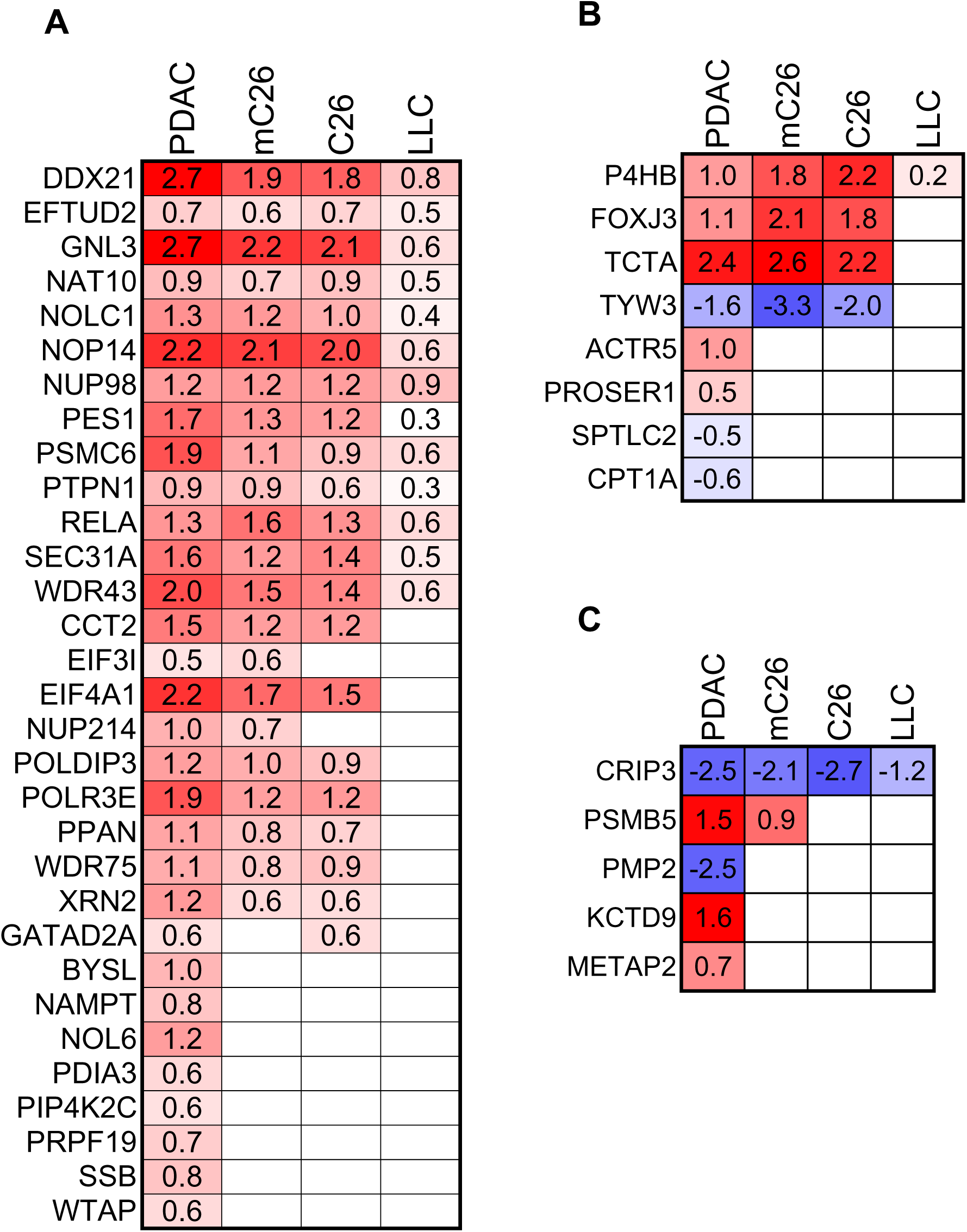
(A) Hub genes identified from humans mapped across different experimental models of cachexia. (B) Mapping of genes identified from machine learning analysis for one-month weight change with experimental models. (C) Mapping of genes identified from machine learning analysis for six months weight change with experimental models. Red and blue indicate upregulated and downregulated genes, respectively.

## Discussion

The current study has systematically profiled the mRNA and miRNA profiles from muscle biopsies of patients with cachexia. To the best of our knowledge, this is the single largest study to profile transcriptome and miRNAs from the same samples using single cancer type to understand the human biology of pancreatic cancer cachexia. This is the first human study in cachexia to comprehensively show the sex-specific differences at the molecular level for PDAC cachexia. Although several common pathways were identified between sexes, the genes involved in those were predominantly different. While common miRNAs and pathways were identified between the two sexes, the gene targets were found to be different, suggesting that sex-specific expression and regulation can be observed at various molecular levels. Further, gene network analysis identified significant sex-specific network modules associated with one-month weight loss compared to the traditional six-month weight loss, indicating that gene changes are more dynamic early in the disease progression. The gene network analyses identified several nodal molecules and their role in cachexia needs to be elucidated further. Sex-specific genes and miRNAs were identified as predictors of weight loss using machine learning approach. Finally, the data generated from same muscle biopsies would serve as an excellent resource to generate new research questions and to compare the results obtained from experimental models.

Sex differences at the molecular level is known for years. However, its importance in physiology and pathology is gaining more attention in recent years. It is well known in cancer biology that sexual dimorphic patterns exist at every level including DNA methylation, mRNA, and miRNA^35, 36^. Similar patterns were observed in our study where minimal overlap was found between sexes for both mRNAs and miRNAs. Network analysis identified several significant modules when sex was considered as a variable, which led us to perform a sex stratified WGCNA. Although several common pathways were identified, the genes involved in activating those pathways were predominantly different. For example, one of the well-known pathways often studied in different experimental models of cachexia is the ubiquitin proteasome pathway, where most of the genes were unique between sexes. Studies have shown that females have higher proteasome activity than males ^37^. Another study shows differing responses to FoxO3a/ ubiquitin proteasome pathway after hindlimb unloading in rats^38^. Similar inferences can be extended to other well-known common pathways, such as mTOR signaling and hedgehog signaling, among others.

Traditionally, a six-month weight loss is used as a part of the classical cachexia definition in classifying patients ^2^. The challenge that accompanies looking at six months of weight loss is that most of the patients, based on the cancer type, would be at the severe to refractory cachexia stage. The muscle from experimental models of cachexia is collected during the active phase of muscle wasting, which may or may not occur in patients at the time of biopsy procurement. As collection of multiple muscle biopsies at different stages of cancer is not feasible in humans due to the invasive nature of the procedure, we utilized the one-month weight loss information in WGCNA analysis to understand the early changes in muscle wasting. Several significant modules at one month weight change in overall and sex-specific analysis identified classical cachexia pathways. This includes ubiquitin proteasome, FoxO signaling, PI3K/AKT signaling^1, 26^ in males and unknown pathways such as Arf6 signaling, Sphingosine-1 phosphate pathway and VEGFR1/VEGFR2 pathway. No significant modules were identified in the six-month weight loss and weight loss grade in males. Similarly, in females, several significant modules were identified in one-month weight change and pathways related to cell cycle, including cell cycle checkpoint, apoptosis, DNA repair pathway, meiotic synapsis, hedgehog signaling, mTOR signaling, among others. Indeed, several of these pathways were also reported in experimental models of cachexia, predominantly using male mice only. Based on these findings, it could be surmised that (i) the early weight loss could be more dynamic in capturing several known signaling pathways associated with cachexia and (ii) there is an urgent need to investigate the mechanistic basis of how several of these well-known pathways would be dysregulated in female models of cachexia.

Hub genes identified from WGCNA are, in general, considered to be functionally important genes with significant impact on the disease. Several studies have experimentally validated the role of hub genes in major depressive disorder^39^. While it is possible to characterize all the identified hub genes, a better curation may aid in narrowing down the list of hub genes for further characterization. One of the approaches we adopted was to compare the hub genes and those obtained from machine learning with different animal models of cancer-associated cachexia. Thirteen genes were common between hub genes identified in humans and experimental models. Apart from RELA, the role of other genes is yet to be characterized in cachexia ^34^. While studies have identified common drivers of muscle wasting across metabolic conditions ^40^, it remains to be seen whether any of these 13 could be functionally important across the cachexia models. From the machine learning approach, P4HB and CRIP3 were the common genes identified from one month and six months of weight loss, respectively. A recent study showed that extracellular vesicles from oesophageal squamous cell carcinoma containing P4HB cause apoptosis through ubiquitin-proteasome pathways ^8^. The role of CRIP3 in muscle wasting remains to be investigated. A similar systematic approach in identifying common genes between humans and mice would provide a comprehensive set of genes that can be functionally characterized and investigated for therapeutic potential. As well, while studying differentially expressed genes is a well-established norm, identifying genes through WGCNA and machine learning approaches can complement the traditional approach.

Thus far, there are two studies that has profiled miRNAs from human skeletal muscle which provided insights on how miRNAs can act as biomarkers for cancer cachexia ^19, 21^. Those studies used muscle from different cancer types and because of the limited sample size, sex-specific analysis was not performed. Both challenges are addressed in the current study, where we observed sex-specific miRNA expression. To the best of our knowledge, this is the first study to profile miRNAs in a sex-specific manner comprehensively. From our *in silico* analysis, we observed that the common miRNAs between males and females regulate genes associated with ketogenesis, tryptophan, and ceramide signaling, among others. Studies suggest that tryptophan catabolism triggered by inflammation has been linked to anemia, which is often observed in patients with PDAC ^41, 42^. Further studies are required to better understand their association to muscle wasting. Recent studies have shown that high levels of ceramides may contribute to muscle wasting by inhibiting anabolic signals in experimental and human models of cachexia ^43^. As miRNAs regulate the genes involved in these pathways, it remains to be tested if modulating those miRNAs can alleviate muscle wasting. When we used the male and female common pathways, several unique and common miRNAs were identified. When we restricted the analysis to the common miRNAs, their gene targets were predominantly different in males and females. For ease of explanation, protein ubiquitination was observed in both males and females, and it shared several common miRNAs. However, their gene targets were predominantly different indicating that the same miRNAs can modulate the same pathway by regulating different genes. Similar trend was observed in other common pathways. These data suggest that miRNAs can regulate the pathways in a sex-specific manner. All these results from gene expression and miRNAs indicate the importance of considering sex as a variable to understand the pathophysiology of cachexia and eventually develop sex-specific therapeutics for better outcomes.

Overall, in addition to providing several insights, ranging from sex-specific expression differences to the use of gene co-expression analysis to identify potential driver molecules associated with muscle wasting, we have also catalogued the list of genes and miRNAs associated with PDAC cachexia. While this is the first step in generating cancer-specific datasets to understand the molecular underpinnings of cachexia in humans, several such efforts are required to further understand the complexity involved in muscle wasting. For example, the same samples can be used to generate the methylome, which may help in understanding the additional regulatory events in muscle wasting. Similar efforts are required for other cancer types, which can eventually be converted into a user-friendly database that may aid in generating more intriguing research questions for discovery purposes.

## Disclosures

TAZ has been compensated for consulting work on cancer cachexia, has carried out sponsored research for Leap Therapeutics, and was/is a member of the Scientific Advisory Board of Emmyon, Inc. and PeleOs. However, none of these financial relationships concern the research presented here. The authors declare no further conflicts of interest.

## Data deposition accession number

pending.

## CRediT Authorship contributions

Ashok Narasimhan, PhD (Conceptualization: Equal, Data Curation: Supporting, Formal analysis: Lead, Methodology: Equal, Visualization: Lead, Writing-original draft: Lead)

Xiaoling Zhong, PhD (Methodology: Supporting, Writing-review and editing: Supporting) Brittany R. Counts, PhD (Formal analysis: Supporting; Data Curation: Equal)

Andrew Young, MS (Data Curation: Equal)

Sha Cao, PhD (Methodology: Equal, Formal analysis: Equal, Writing-review and editing: Supporting)

Jun Wan, PhD (Data Curation: Equal, Writing-review and editing-Supporting) Sheng Liu, PhD (Data Curation: Equal, Writing-review and editing-Supporting)

Leonidas G. Koniaris, MD (Conceptualization: Supporting, Data Curation: Supporting, Writing-review and editing-Equal)

Teresa A. Zimmers, PhD (Conceptualization: Lead, Data Curation: Equal, Formal analysis: Equal, Funding acquisition: Lead, Resources: Lead, Supervision: Lead, Writing-original draft: Supporting, Writing – review & editing-Lead)

## Data Transparency statement

All the data associated with the manuscript including the anonymized clinical information will be submitted to Gene Expression Omnibus database.

## Data Availability

All data produced in the present study will be available online in GEO upon publication.

## Acknowledgments

This work was supported in part by grants from the Lustgarten Foundation (TAZ), the National Institutes of Health, including P01 CA236778-01A1 (SC, LGK, TAZ), R01CA257452 (TAZ), T32 CA254888 (BRC), NIH/NCI 1L70CA284463-01 (BRC), the Knight Cancer Institute (NIH grant P30 CA069533), the U.S. Department of Veterans Affairs grant I01CX002046 (TAZ), and the Brenden Colson Center for Pancreatic Care at Oregon Health & Sciences University.

Specimen collection and histology were supported by the Biospecimen Collection and Banking Core of the Indiana University Simon Comprehensive Cancer Center (IUSCCC) (NIH grant P30CA082709). Bioinformatics analysis was supported by the Walther Cancer Foundation, the Purdue University Center for Cancer Research (grant P30CA023168) and the IUSCCC. Additional support was provided by the Adams County Cancer Coalition (TAZ).

## Table legends

**Table 1**: Patient demographics: Values are represented as mean ± standard deviation. *= t-test, †= chi-square test, ‡= Fisher’s exact test. CT information was not available for 2 control subjects and 8 PDAC subjects. Information on 1 MOWLG was not available in one control and 6 MOWLG on two controls and 1 PDAC subject.

MOWLG – Month weight loss grade, MWC – Month weight change

**Table 2**: Machine learning analysis – list of sex-specific mRNAs and miRNAs.

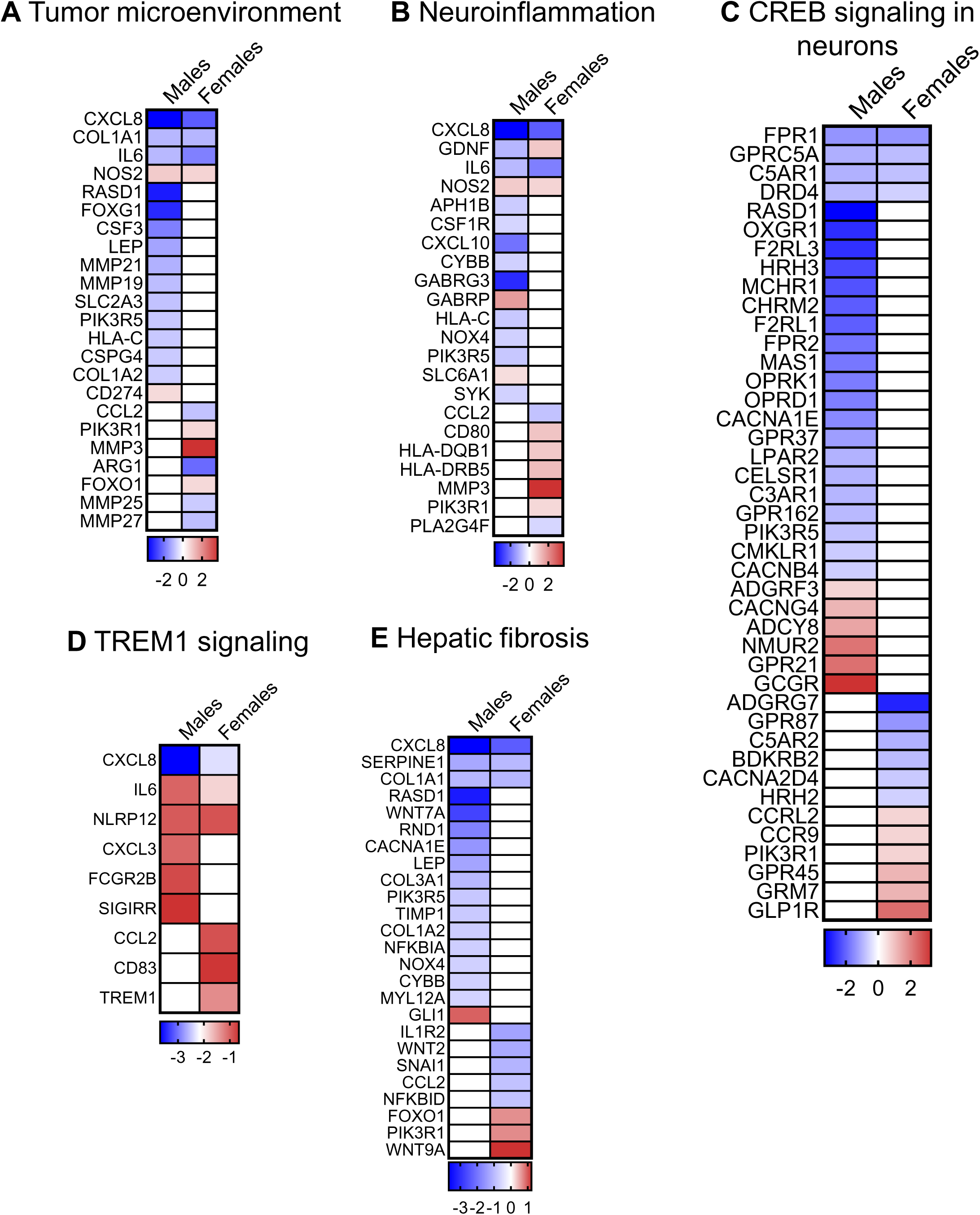
Figure S1:

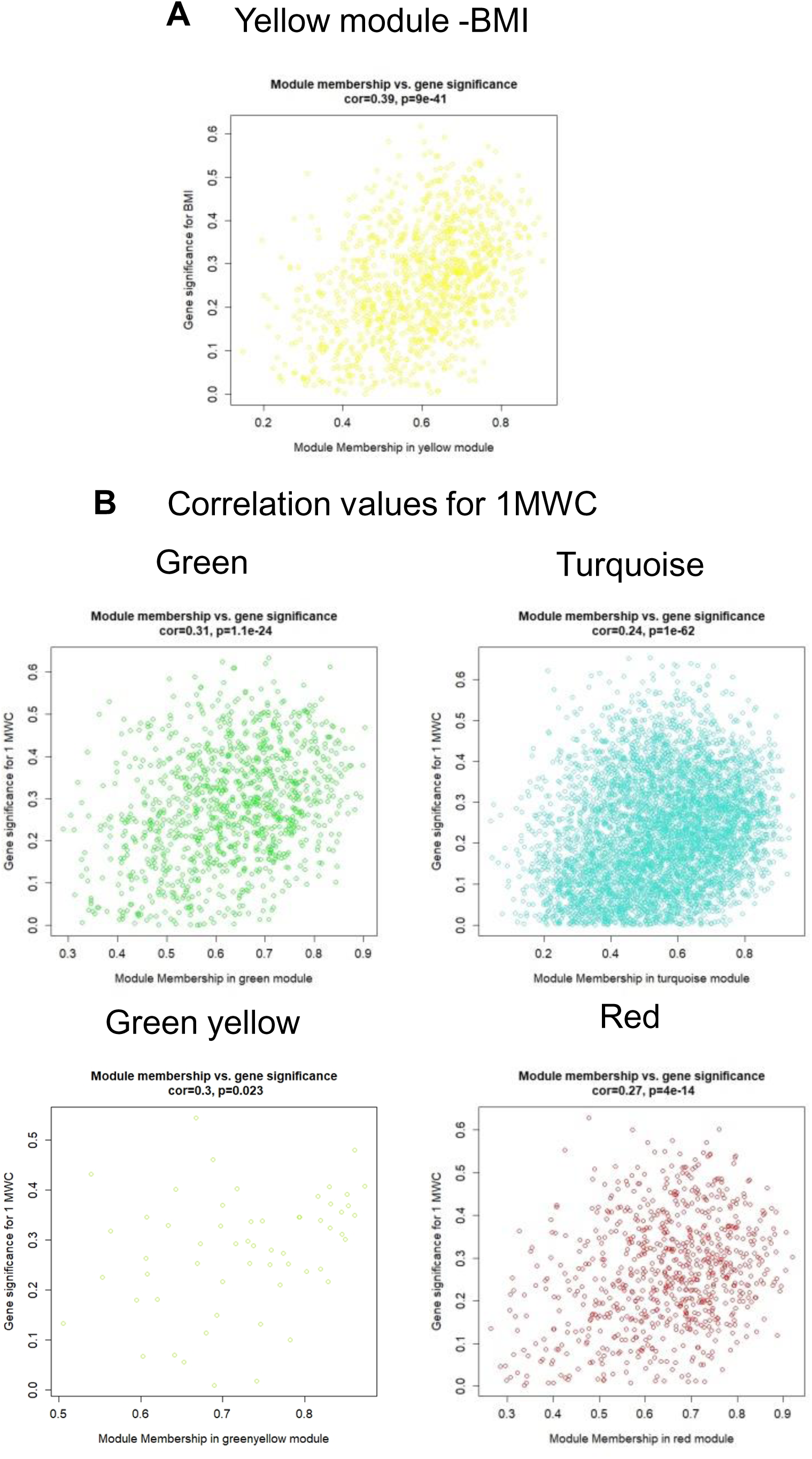
Figure S2:

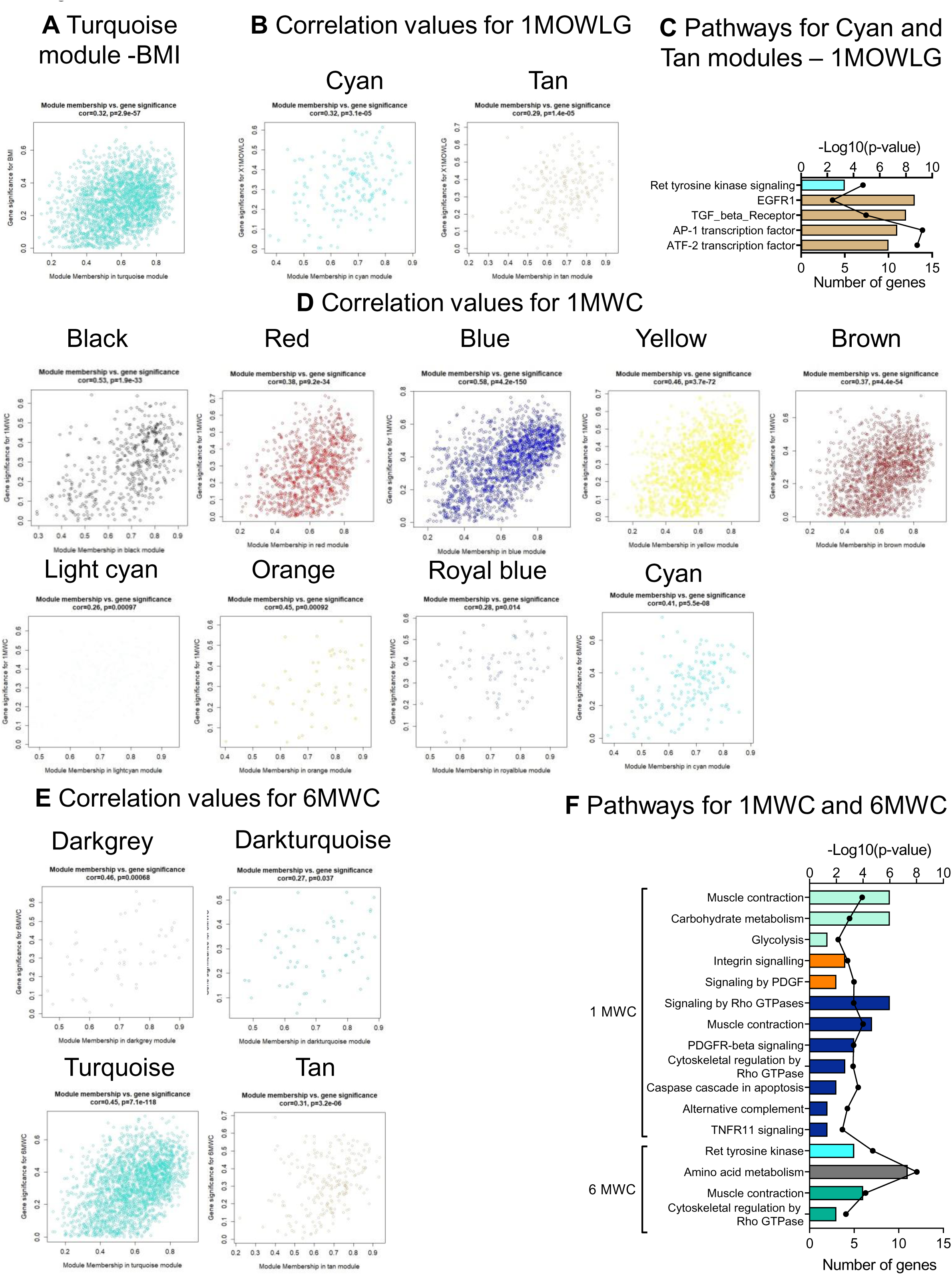
Figure S3:

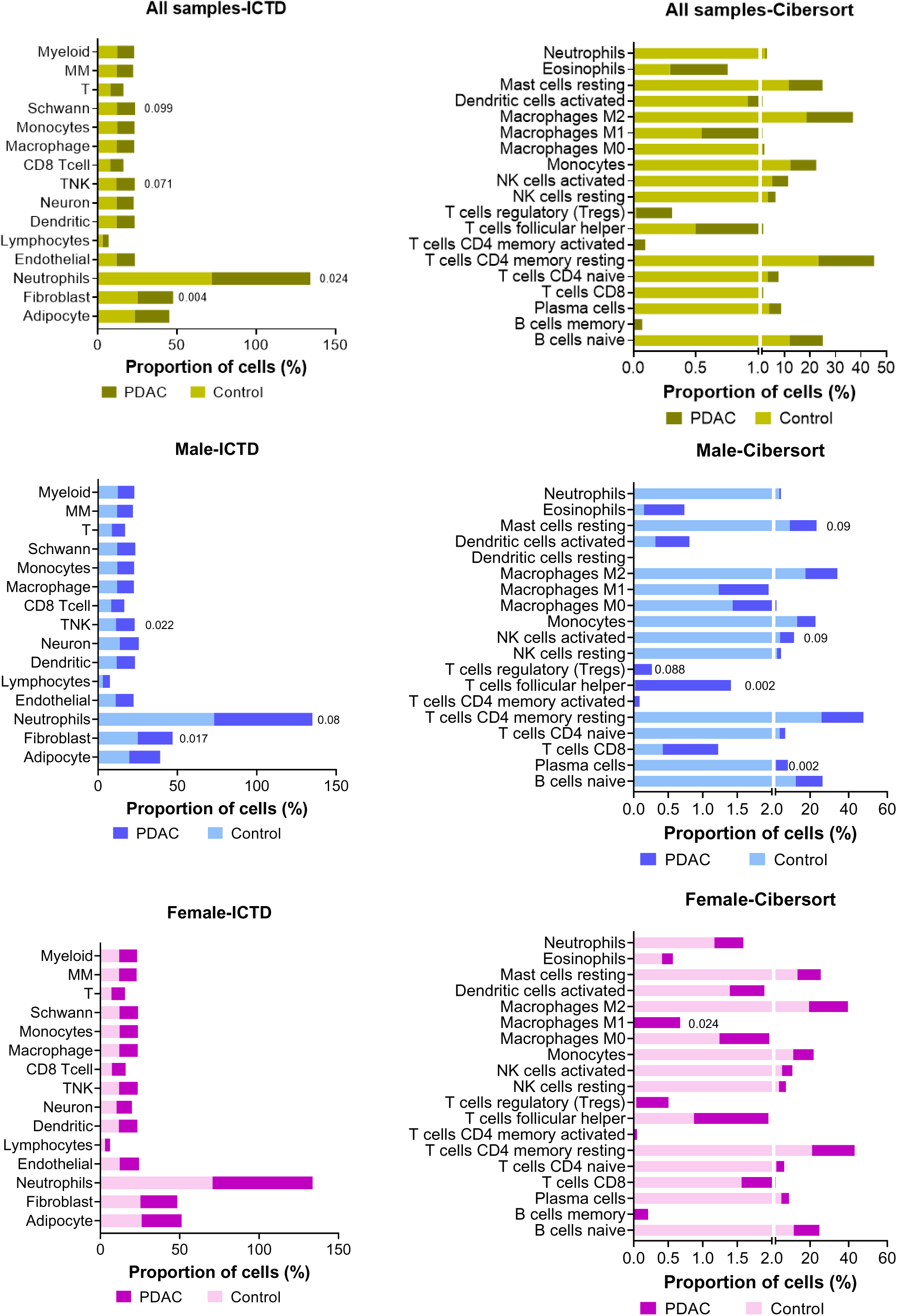
Figure S4:

## Notes

### Author Declarations

The IRB of Indiana University gave ethical approval for this work.

